# SARS-CoV-2 detection is independent of microbiome composition on surfaces in a major Ontario hospital

**DOI:** 10.1101/2025.06.03.25328878

**Authors:** Nikhil A. George, Lauren Bradford, Aaron Hinz, Marita El Kadi, Lydia Xing, Evgueni Doukhanine, Derek R. MacFadden, Caroline Nott, Michael Fralick, Rees Kassen, Alex Wong, Laura A. Hug

## Abstract

The SARS-CoV-2 pandemic has resulted in considerable mortality in hospital settings. Built environment surveillance can provide a non-invasive indicator of SARS-CoV-2 status in hospitals, but we have a limited understanding of SARS-CoV-2’s microbial co-associations in the built environment, including any potential co-occurrence dynamics with pathogenic and antimicrobial-resistant microorganisms. Here we examine the microbial communities on floors and elevator buttons across several locations in two major tertiary-care Ontario hospitals during a surge in SARS-CoV-2 cases in 2020. Total microbial community composition, prevalence and type of detected antimicrobial resistance genes, and virulence factor distributions were governed by sample source rather than SARS-CoV-2 detection status. Fifteen microorganisms were identified as indicator species associated with positive SARS-CoV-2 signal, including three opportunistic pathogens (i.e., two *Corynebacterium* sp. and a *Sutterella* sp). Key clinically relevant antimicrobial resistance genes showed varying prevalence across sites within the hospital, suggesting that our workflow could inform resistance burden in hospitals. Overall, these results indicate limited or only weak interactions between microbiome composition and SARS-CoV-2 detection status in the hospital built environment.

## Introduction

Since its identification in December 2019, severe acute respiratory syndrome coronavirus-2 (SARS-CoV-2, the etiological agent of COVID-19) has caused a global pandemic, with over 777 million cases and associations with over 7 million deaths (as of February 2025, World Health Organization, https://covid19.who.int). Human-to-human transmission through aerosolization of the virus is the primary route of transmission [1]. Outbreaks of COVID-19 remain common in both hospitals and long-term care homes worldwide, leaving both patients and healthcare workers at high risk [2]. Monitoring of viral detection and burden in the built environment can provide information on disease burden and infection risk [3,4].

Recent research has detected SARS-CoV-2 on various hospital surfaces, with detection on floors being most frequent [5–8]. Detection frequency has been strongly associated with infection burden within the hospital [9]. Although instances of fomite transmission are limited [1], SARS-CoV-2 detection on fomites in hospitals has been predicted by bacterial signatures [10], suggesting that some bacteria, and possibly other microorganisms, may influence SARS-CoV-2 transmission and propagation. Further, bacteria and other microbes can metabolize and transform a variety of chemicals in their environment, including disinfectants [11,12], potentially neutralizing or attenuating the effects of these agents against viruses.

Surface sampling for SARS-CoV-2 has advanced our understanding of where the virus can be detected within a built environment, and has yielded new surveillance methods for hospital COVID-19 cases [6,9]. There have been limited examinations of microbial co-associations with SARS-CoV-2 in hospital environments to date. One large-scale 16S rRNA amplicon sequencing survey examining these co-associations across patients, health care providers, and the built environment identified that floor samples carried the highest microbial biomass [10]. This survey also identified a single amplicon sequence variant, from the genus *Rothia*, that was strongly associated with SARS-CoV-2 detection. Pochtovyi and colleagues surveyed a Moscow hospital, swabbing floors, door handles, and electronics in several wards and the intensive care unit (ICU) to determine prevalence of six pathogens with expected co-occurrence with SARS-CoV-2 infection [13]. Targeted PCR identified at least one of these six pathogens of interest from each built environment sample. 16S rRNA amplicon sequencing identified more diverse communities on floors compared to other surfaces, and identified several lineages that were characteristic for the different units (ICU versus Respiratory Infections Department) [13]. These studies indicate microbiome co-occurrence with SARS-CoV-2 can be detected from the built environment and may be informative for infection control. However, amplicon sequencing cannot determine microbiome features with clinical relevance, such as antimicrobial resistance genes (ARGs), leaving open questions as to the nature of the built environment microbiome in relation to SARS-CoV-2.

Here, we conduct deep metagenomic sequencing on biomass collected from swabs of floors and elevator buttons to identify microbial community characteristics associated with SARS-CoV-2 detection and prevalence within a hospital environment. We assess the gene complement of antimicrobial resistance genes, viral prevalence, and virulence factors to determine the interactions of SARS-CoV-2 with hospital microbiota.

## Materials and methods

### Sample collection

Sample collection and SARS-CoV-2 detection was previously described [7]. In brief, samples were obtained over a 10-week period (September 28th, 2020 to December 6th, 2020) from two tertiary-care, academic hospital campuses from Ottawa, Ontario, Canada (Campus A and Campus B, totaling over 1100 beds), encompassing 8 different sites (ICU, elevators, parking garage, and both COVID and non-COVID wards). Samples for metagenomics were selected from this larger-scale study which specifically and exclusively examined the spatio-temporal dynamics of SARS-CoV-2 detection [7]. Surfaces were sampled with the P-208 Environmental Surface Collection Prototype kit (DNA Genotek, Ottawa, ON). This included swabbing a surface and then submerging the swab in a nucleic acid stabilization solution for subsequent storage at ambient temperature for up to one month. Before processing, control samples were generated through a 10-20 second exposure of a swab to the air of a location without any swabbing. The MagMAX Viral/Pathogen II (MVP II) Nucleic Acid Isolation Kit (Thermo Fisher Scientific, Waltham, MA) was used for total nucleic acid extraction. Extracted RNA was used as input to quantitative reverse-transcriptase polymerase chain reactions (RT-qPCRs) to detect SARS-CoV-2.

Samples for metagenomics were chosen from three wards with SARS-CoV-2 detection and three wards with no SARS-CoV-2 detection over the course of the larger-scale survey [7]. Samples were selected to allow a comparison of locations (wards, ICU, parking garage), site types (floors versus elevator buttons), and SARS-CoV-2 detection status as previously determined through RT-qPCR [7]. Sample information is available in Supplemental Table 1.

### Sequencing and quality control

Thirty-eight samples were submitted to the CAGEF DNA sequencing facility (Toronto, Canada). Libraries were constructed with the Illumina Nextera Flex kit and then converted into DNA nanoballs (DNBs) for sequencing on the MGI G400 using the MGI FCS PE150 sequencing kit. Adapter removal and quality filtering for sequenced reads was performed using Trimmomatic [14] and sickle [15].

### Taxonomic assignment of reads

Assembly attempts were unsuccessful, yielding small numbers of short, low-coverage contigs, so we adopted a read-based approach. Quality-controlled reads were taxonomically assigned using Kraken2 [16] with the flags “--confidence” (set to 0.65) and “--report-zero-counts,” and the NCBI RefSeq Complete V205 500GB Kraken2 database [17], which is a substantially more complete database than the default distribution.

Bracken2 [18] was used to refine Kraken2’s estimates of reads assigned to each taxon and resolve potential misclassifications. Exploratory data analyses of Kraken2 and Bracken2 output were performed using PAVIAN [19].

### Microbial community analyses

Microbial community analyses were conducted using the R Statistical Software v4.1.1 [20]. Data handling was accomplished using the phyloseq v.1.48.0 [21], tidyverse v.2.0.0 [22], and reshape2 v.1.4.4 [23] packages. Data was visualized using the dplyr v.1.1.4 [24] and ggplot2 v.3.5.1 [25] packages.

Taxonomic assignments for reads were imported from Bracken2 output files, and reads associated with the genus *Homo* were removed from all further analyses, in accordance with the ethics approval/REB associated with the original sampling [7]. Reads associated with the genus *Homo* were also removed from the raw read files deposited to the NCBI using the extract_kraken_reads.py script from the KrakenTools package [26]. The remaining Eukaryotic reads were very patchily distributed and primarily associated with plant matter (*e.g.*, *Brassica*). All non-*Homo* Eukaryotic reads were subsequently removed from the datasets prior to microbial community composition analyses but were kept for data deposition to NCBI.

Microbial community compositions were compared using Principal Coordinates Analysis (PCoA) across SARS-CoV-2 detection status and location source (Elevator, Floor) using phyloseq’s ordinate function and a Bray-Curtis dissimilarity statistic [27]. The Shannon alpha diversity metric was calculated for all samples and compared across SARS-CoV-2 detection status and source [28]. Indicator species analyses were conducted using the package indicspecies v.1.7.15 [29] to identify key taxa specifically associated with SARS-CoV-2 positive or negative detection status.

Viral reads were subset and analyzed as a separate community, including PCoA, alpha diversity measures, and indicator species analysis as described for the full dataset above.

### Virulence Factor and Antimicrobial Resistance Gene Predictions

All reads were mapped to the core Comprehensive Antibiotic Resistance Database (CARD) v3.2.7 (downloaded June 12, 2023) [30] and core Virulence Factor Database (VFDB) (downloaded July 05, 2023) [31]. RGI v6.0.0 [30] was used for additional mapping to the core CARD. Predicted virulence factor and antimicrobial resistance gene prevalence and sample ordinations based on annotation categories were generated and visualized in R. Prevalence was normalized to reads per million to enable comparisons across samples.

Clinically relevant ARGs were selected (*blaCTX-M, blaKPC, blaNDM-1, blaOXA, blaVIM, mecA, mecC, vanA, vanB*), and annotations were manually harmonized to allow filtering of the datasets by this subset of ARGs. Kruskal-Wallis tests were used to determine if ARGs associated significantly across SARS-CoV-2 positivity status, sample source, or unit (ICU and Ward #) [32]. Pairwise comparisons between ARG abundance profiles were conducted using the Wilcoxon rank sum test with continuity correction [33], with a Benjamini-Hochberg adjustment to p-values to account for multiple tests [34].

## Results and Discussion

Metagenomes were generated for a total of 38 samples (of a total 963 swabs, 4.0%, Supplemental Table 1, [7]). Sample read counts ranged from 31,839,940 to 444,624,914 (median 104,749,332; control sample 3,902,008). Following taxonomic assignment of reads by Kraken2 and Bracken [16,18], all reads associated with the genus *Homo* were removed to meet study ethics requirements, removing 0.66-65.30% of the trimmed and quality-filtered reads (average 13.5%). Eukaryote-free datasets contained 1,909,955 to 39,486,112 reads classified to at least the domain level (median 7,344,826; control sample 590,963). The control sample was not expected to be aseptic, as it was an air swab rather than a sterile control.

### Bacterial reads dominate Eukarya-free datasets

Samples varied in their bacterial, archaeal, and viral composition, with the Bacteria most abundant across all samples (Supplemental Figure 1). Samples were dominated by reads from four phyla: Actinomycetota (53.8%), Pseudomonadota (24.0%), Bacillota (16.7%), and Bacteroidota (4.4%) (Figure 1A). Reads from the other 34 detected phyla and reads unclassified at the phylum level comprised only 1.1% of the total reads (Figure 1A). Only the Cyanobacteria and Verrucomicrobiota were also present above 1% abundance in any individual sample, with Cyanobacteria above 1% in 10 and Verrucomicrobia above 1% in 1 sample(s) respectively. At the genus level, 21 genera had an abundance above 5% in at least one sample (Figure 1B), and 11 genera had an abundance above 1% across all samples. Notably, *Cutibacterium* dominated most samples, with other human-skin associated genera also strongly represented, including *Pseudomonas*, *Corynebacterium*, and *Staphylococcus* [35].

**Figure 1:**
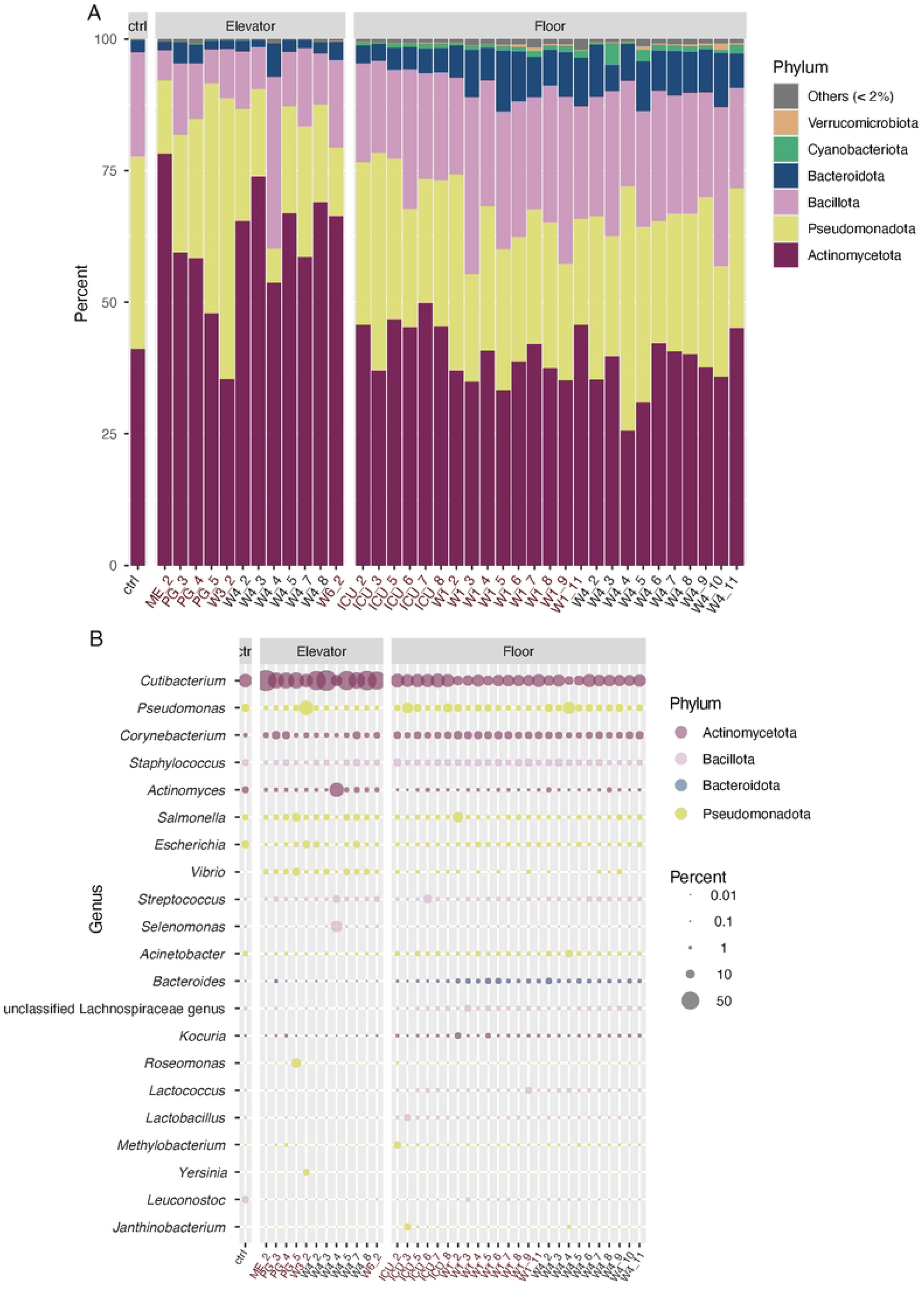
Microbial community composition overview at the (A) phylum and (B) genus levels. In A, all phyla present at or above 1% abundance in at least one sample are depicted as bars, with lower abundance phyla grouped into the “Others” category. In B, only genera present above 5% abundance in at least one sample (21 genera) are included. Bubbles are coloured by phylum and scaled by percent total abundance of the given sample. Sample names follow the convention UNIT_WEEK, as facets depict elevator vs. floor samples. Swabs positive for SARS-CoV-2 are colored red.

Viral communities showed higher variability across samples, with certain viruses identified sporadically at very high relative abundance (Figure 2). Viral groups with consistent presence across hospital samples were from the class Caudoviricetes, the order Zurhausenvirales, and, at a lower abundance, the order Crassvirales (Figure 2A). Looking at specific viruses, three unclassified Caudoviricetes were consistently identified across samples, as was an uncultured human fecal virus (Figure 2B). Two papillomaviruses, one Betapapillomavirus and one human papillomavirus, were also consistently detected across samples, with the human papillomavirus showing an increased abundance in the Ward 6 elevator sample from week 2 (Figure 2B). Other viruses with very high abundances in a single sample were a Lymphocryptovirus humangamma4 (Ward 1 Floor, week 2), a *Streptococcus* phage (ICU floor in week 6), and, at a lower abundance, the Betapapillomavirus described above (parking garage elevators in week 4). It is notable that each of these specific viral spikes in abundance were identified from SARS-CoV-2 positive samples, though none of these were identified as indicator species for SARS-CoV-2 detection (see below).

**Figure 2:**
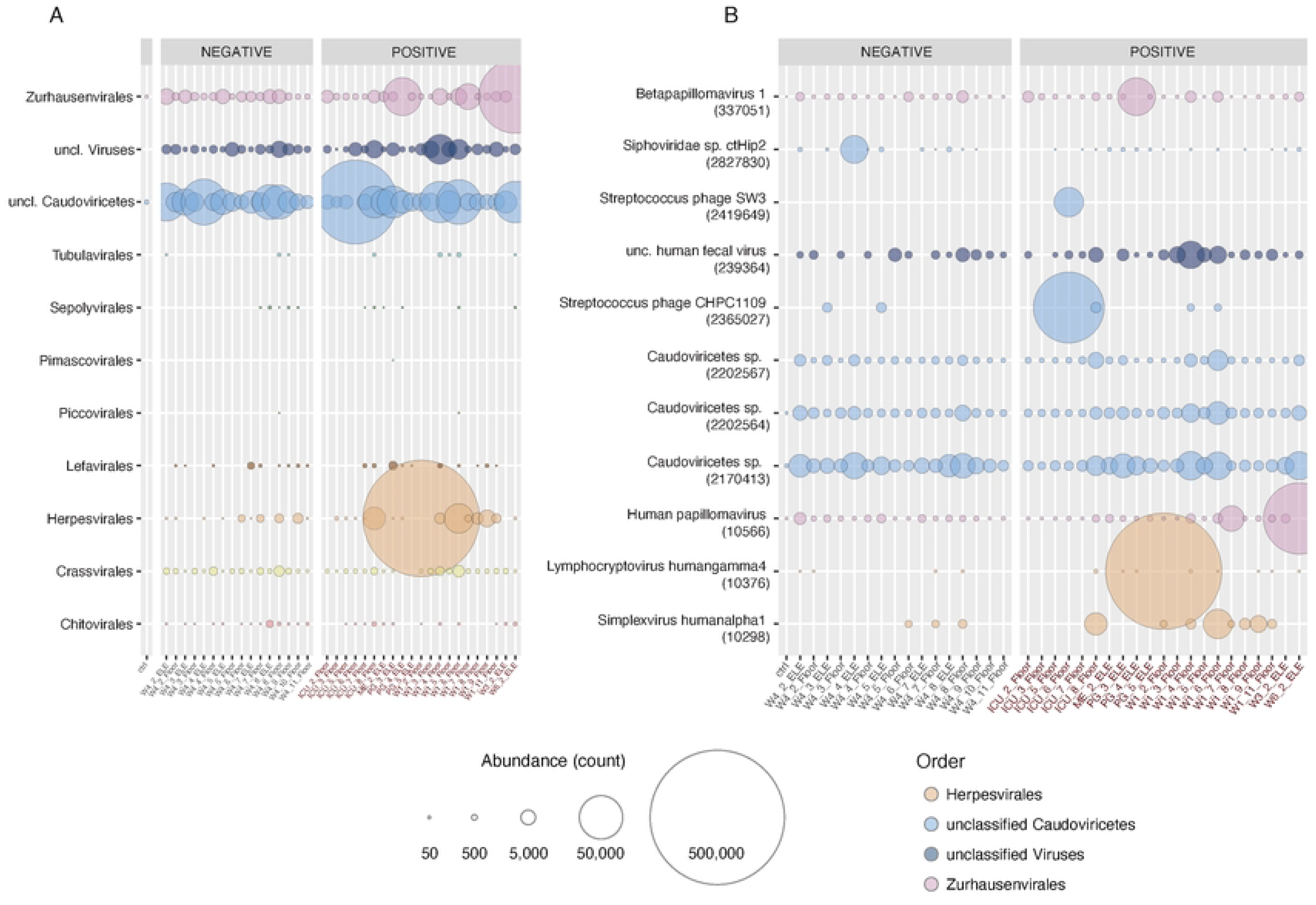
Viral community composition overview at the (A) order and (B) unique virus levels. Bubbles are scaled by normalized read count. In A, all viral orders identified in the data are included in the bubble plot. In B, only viruses identified at over 10,000 reads in at least one sample are depicted. Bubbles are coloured by viral order as in A. Facets represent samples that were negative or positive for SARS-CoV2 detection in a previous study [7]. Sample names follow the convention UNIT_WEEK_SOURCE, with positive SARS-CoV-2 samples colored red as in Figure 1.

The overall Shannon diversity was significantly lower in elevator samples relative to floor samples (Supplemental Figures 2, 3). Shannon diversity across positive and negative SARS-CoV-2 detection status was not significantly different for either Floor or Elevator samples. For viral communities, there was no significant difference in Shannon diversity across Source or SARS-CoV-2 detection status (Supplemental Figure 4).

### Sample composition clusters according to source and not SARS-CoV-2 detection status

Principal coordinates analysis (PCoA) on Eukaryote-free datasets showed grouping based on sample source (*i.e.*, floor, elevator, or control), with limited overlap between floor and elevator samples (Figure 3A). SARS-CoV-2 detection status did not map to clear groups as defined by sample biodiversity. When PCoA groupings were examined by location (ICU, Wards, etc.), samples from the ICU grouped tightly, whereas samples from other sites were more widely spread out (Supplemental Figure 5). When only the viral subset of the datasets was used to generate the PCoA plot, a stronger degree of overlap was observed for floor and elevator samples (Figure 3B), with SARS-CoV-2 positive samples exhibiting more variation on the plot than SARS-CoV-2 negative samples, reflective of the sporadic detections of unique, high-abundance viruses in certain samples (Figure 2).

**Figure 3:**
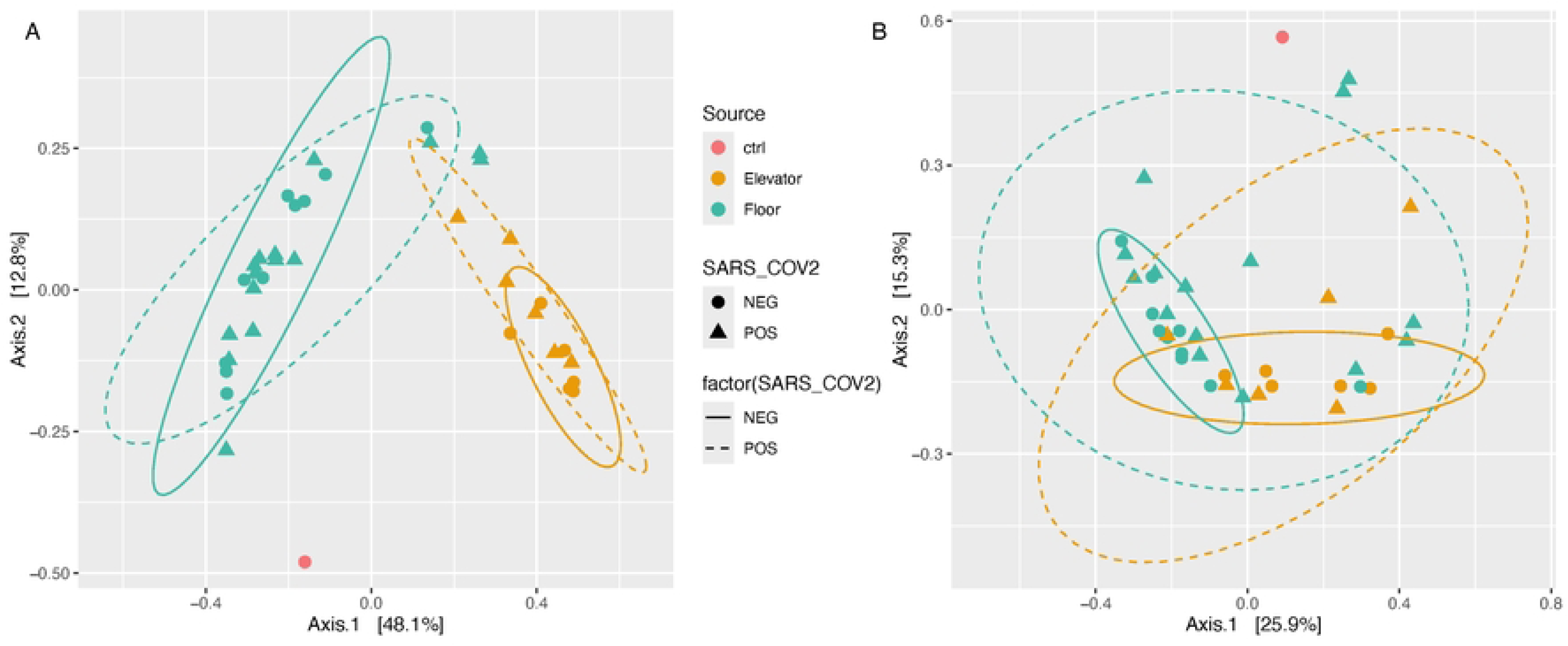
PCoA of sample microbial community composition (Bacteria, Archaea, viruses) (A) and viral diversity only (B), coloured by source type (elevator, floor, and control (ctrl)), with SARS-CoV-2 detection status indicated by shape. Ellipses encompass SARS-CoV-2 positive (dashed) and negative (solid) samples within each site type.

### Bacterial and viral indicator species based on SARS-CoV-2 detection status

While overall microbial community compositions were governed by sample source, multiple taxa showed strong associations with SARS-CoV-2 detection. Fifteen microorganisms showed positive association with SARS-CoV-2 detection, of which the most strongly correlated were a *Sphingobacterium* and a *Lentilactobacillus* species (Figure 4). Eight of the fifteen microorganisms are from the phylum Pseudomonadota. One archaeon, *Halococcus hamelinensis*, was identified as an indicator species for positive SARS-CoV-2 detection. No viruses showed positive correlation with SARS-CoV-2 detection. None of the 15 indicator species are obligate pathogens in humans. Three are opportunistic pathogens of humans: the two *Corynebacterium sp*. and the *Sutterella sp.* KLE1602. These three are also the only indicator species associated with the human microbiome, with *Corynebacterium* a common skin-associated lineage and *Sutterella* commonly found in the human gastrointestinal tract. Of the remaining twelve microorganisms, seven are typically associated with soil and sediment environments, two are psychrotolerant (*Psychrobacter* and *Pontibacter arcticus*) and one is halophilic and associated with hypersaline lakes (*Halococcus hamelinensis*). The *Rickettsia* identified is an endosymbiont of the black-legged tick, which could have medical associations in a hospital environment.

**Figure 4:**
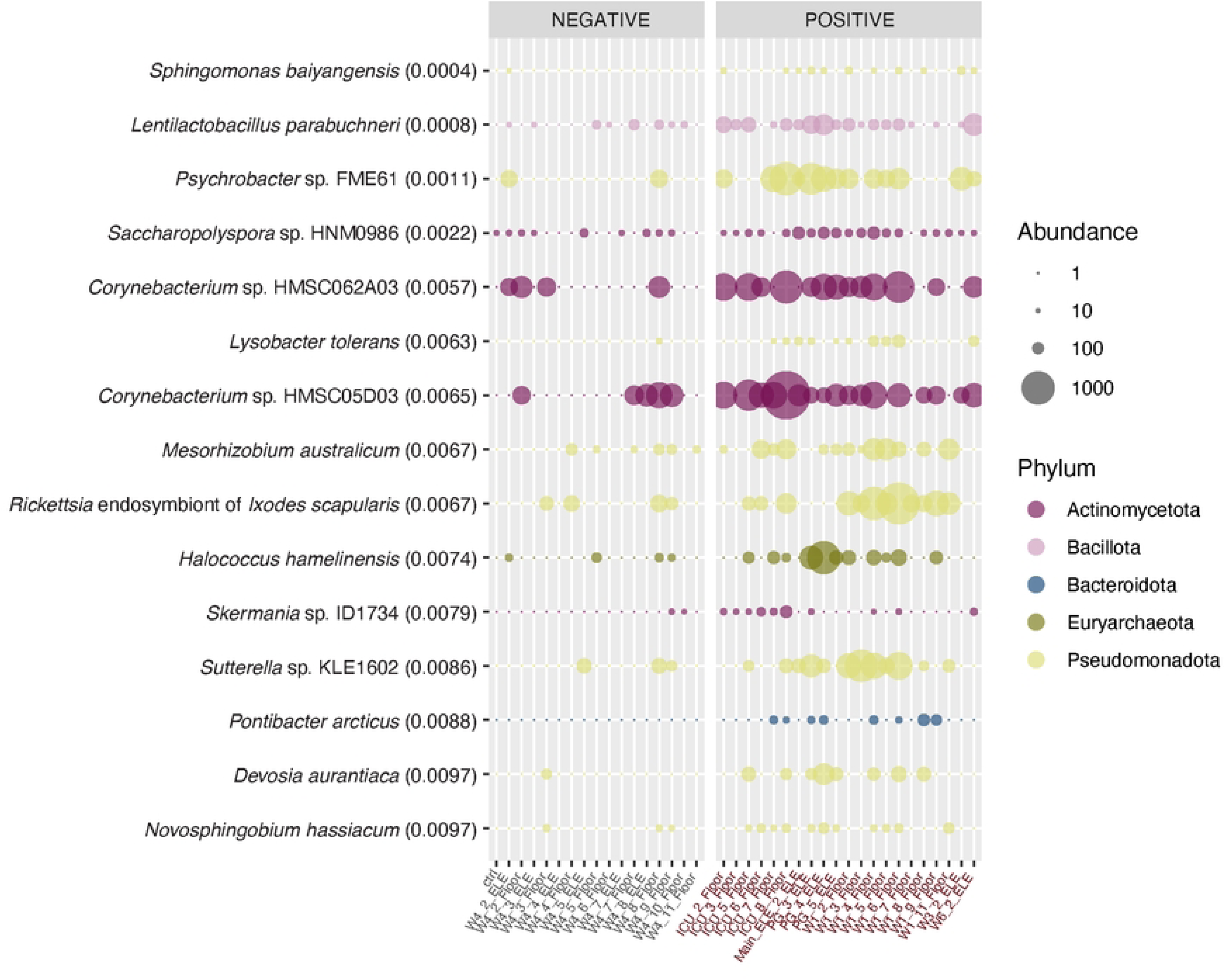
Abundance profiles across samples for organisms identified as significantly positively associated (p < 0.01) with SARS-CoV2 detection by indicator species analysis. Facets represent samples that were negative or positive for SARS-CoV2 detection in a previous study [7]. Bubbles are scaled by abundance as normalized read counts, and coloured by phylum. Organisms are ranked by significance, with their individual p-value (p < 0.01) from the indicator species analysis noted after each organism’s name in brackets. Indicator species for negative SARS-CoV-2 detection status are presented in Supplemental Figure 6.

*Lentilactobacillus parabuchneri* is commonly found in dairy products, and thus its presence may be due to patient, staff, or visitor foodstuffs.

Thirty-six organisms, including one virus, were identified as indicator species associated with negative SARS-CoV-2 detection, demonstrating a negative correlation with SARS-CoV-2 detection (Supplemental Figure 6). The organisms most strongly negatively correlated with SARS-CoV-2 detection were *Microbacterium liquefaciens*, *Salicibibacter halophilus*, and *Tsukamurella tyrosinosolvens* (Supplemental Figure 6). We note that no members of the genus *Rothia* were identified as indicator species in this analysis, contrasting findings from an earlier survey of microbial co-associations with SARS-CoV-2 in a U.S. hospital [10]. This may be due to geographic differences in hospital microbiomes, or differences between study methodologies and analysis tools. Sixteen of thirty-six organisms are from the phylum Pseudomonadota, while ten are members of the Actinomycetota.

### Virulence factor profiles

The distribution of virulence factors within the VFDB core dataset was examined across samples. A high proportional abundance of virulence factors was observed in a single elevator sample (Ward 3, week 2 (W3_2_ELE), Supplemental Figure 7). PCoA analysis revealed that, akin to microbial diversity, sample source – rather than SARS-CoV-2 detection status – governed the observed virulence factor profile distributions. Consistent with its higher overall virulence factor signal, Ward 3’s week 2 elevator sample had the most distinct virulence factor profile relative to other samples (Supplemental Figure 8, upper right).

### Antimicrobial resistance genes

Antimicrobial resistance genes (ARGs) were detected by mapping reads to the Comprehensive Antimicrobial Resistance Database (CARD). In this analysis only, the control sample demonstrated an unexpectedly high proportional abundance of ARGs (Supplemental Figure 9A). The control sample had a much lower overall read count, which may have led to an over-inflation of predicted ARGs when normalized to counts per million reads. The control sample also showed a distinctly different microbial community composition across all metrics, and as such this enrichment in ARGs, if not an analysis artifact, is hypothesized to be due to contamination from DNA extraction and sequencing processing kits, where antimicrobial-resistant organisms were likely used to generate the relevant enzymes. None of the ARGs identified as clinically relevant for further analyses (see below) were detected in the control sample. We removed ARGs associated with the control sample from all samples’ predicted ARGs - resulting in a removal of 121 of the 1,830 identified ARGs (6.6%; Supplemental Figure 9B). Two elevator samples, Ward 3 week 2 and Ward 6 week 2, had the greatest proportional abundance of ARGs. PCoA analysis showed floor samples and elevator samples had distinct ARG profiles; however, floor samples contained higher intra-group similarity in ARG profiles relative to elevator samples, and elevator samples positive for SARS-CoV-2 detection specifically drove this intra-group variability (Supplemental Figure 10).

ARGs were categorized by CARD Resistance Mechanisms, which showed similar profiles across Source and SARS-CoV-2 detection variables. Antibiotic efflux was the most abundant resistance mechanism, with antibiotic inactivation and antibiotic target alteration also strongly represented (Supplemental Figure 11). By Drug Class, resistance to tetracycline, aminoglycoside, mupirocin-like, and peptide antibiotics showed elevated prevalence. The most abundant type of antibiotic resistance was multidrug resistances, where multiple antibiotic classes were targeted by the same resistance mechanism (Supplemental Figure 12). Between floors and elevators, there were differences in prevalence of certain antibiotics: rifamycin and glycopeptide resistance were elevated in floor samples, whereas fluoroquinolone antibiotic resistance was at a higher prevalence in elevator samples (Supplemental Figure 12). Given the importance of disinfection within the hospital built environment, we were interested in the prevalence of resistance to disinfecting agents, which was a stand-alone Drug Class as well as being included in several multidrug resistance Drug Classes. Resistance to disinfection agents represented 8.1% of ARG-associated reads, with an average prevalence of 19.35 reads per million reads (Supplemental Table 2). Elevator buttons carried a slightly higher prevalence than floors, but there were no significant differences by Source (Elevators: mean 20.8 +/-17.8 rpm; Floors: mean 18.7 +/-5.6 rpm; p=0.69). Similarly, there was no significant difference between SARS-CoV-2 positive and negative samples (Positive: 20.3 +/-14.4 rpm; Negative: mean 18.0 +/-4.2 rpm; p=0.48). Higher variance in the Elevators and SARS-CoV-2 positive statistics were driven by the Ward 3 week 2 elevator sample’s higher ARG prevalence.

We next examined the distribution of specific clinically relevant ARGs known to confer resistance to beta-lactams, carbapenems, and glycopeptides: *blaCTX-M, blaKPC, blaNDM-1, blaOXA, blaVIM, mecA, mecC, vanA, and vanB*. The relationship between ARG abundance and Unit was not significant for these ARGs of interest (Supplemental Table 3). A Kruskal-Wallis test examining ARG abundance across SARS-CoV-2 detection status was not significant (p = 0.31). Comparison of the selected ARGs’ abundances across sample source (elevator button vs. floor) was significant (p < 0.0092), driven by *vanA* showing a significantly higher prevalence on floors compared to elevator (p < 1.74 x10^-5^; Supplemental Figure 13, Supplemental Table 3). This indicates ARG burden is not correlated with SARS-CoV-2 incidence across Units, and that floors likely remain the most effective sampling location for hospital-based built environment surveillance.

A Kruskal-Wallis test based on ARG abundance across samples identified a significant difference (p < 2.2x10^-16^) by abundance of these ARGs. A pairwise Wilcoxon rank sum test using a Benjamini-Hochberg correction for multiple tests identified four distinct groups based on ARG abundance profiles: *blaNDM-1, blaVIM, mecC,* and *vanB* (Group 1, very low abundance); *blaCTX-M* and *blaKPC* (Group 2, low abundance); *vanA* alone (Group 3, sporadic high abundance), and *blaOXA* with *mecA* (Group 4, high abundance) (Figure 5, Supplemental Table 4).

**Figure 5:**
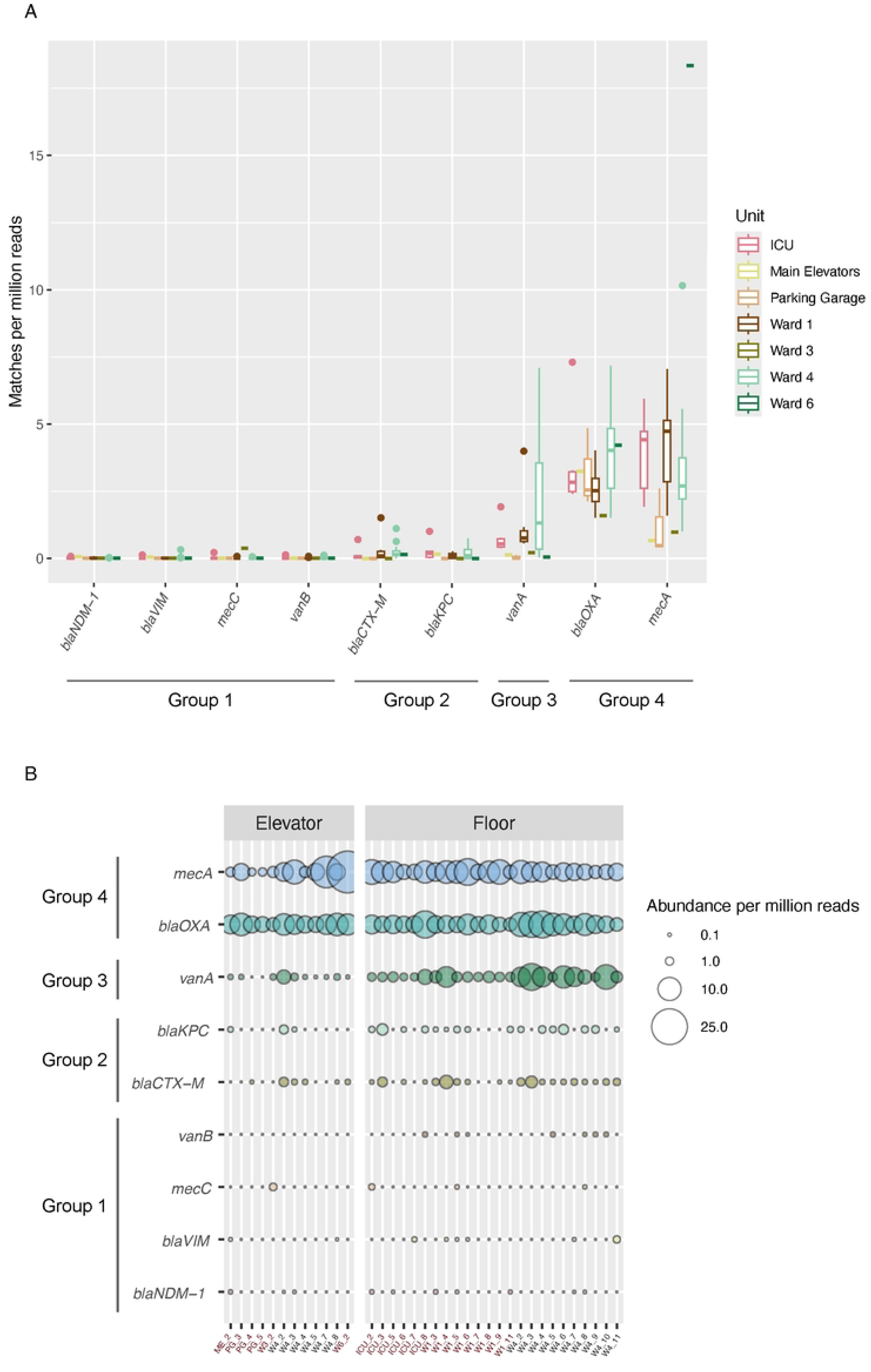
A: ARGs of interest abundance across hospital Units, organized by groups identified through a pairwise Wilcoxon rank sum test: Group 1 – low abundance; Group 2 – very low abundance; Group 3 – sporadic high abundance; and Group 4 – high abundance. Box plots indicate the first and third quartile of results with the thick horizontal bar indicating the median. Whiskers identify the range of the data, with outlier data points plotted. **B:** Bubble plot showing ARGs of interest abundance across samples. Sample names follow the convention UNIT_WEEK, as facets depict elevator vs. floor samples. Swabs positive for SARS-CoV-2 are colored red.

Among the clinically relevant ARGs of interest, the most abundant were *blaOXA, mecA, and vanA*. *blaOXA* and *mecA* were identified at all seven Units, while *vanA* was specifically detected in the ICU, Ward 1, and Ward 4. The three higher-abundance ARGs profiles changed in abundance across Units and across weeks (Figure 5), indicating this built-environment monitoring protocol does show variation in the ARG burden on hospital surfaces. Additional work is required to determine if this protocol provides insight into the ARG burden within the patients on these wards, which would extend the utility of built environment surveillance.

## Conclusions

Metagenomic sequencing of swabs from hospital floors and elevator buttons identified a diverse microbial community. Bacterial and archaeal community membership was more stable across samples than the viral community, which was defined by sporadic viruses with elevated abundances. These included viruses with microbial hosts as well as human viruses (e.g., papilloma viruses and a simplex virus). Community diversity was more strongly defined by sample source (floors vs. elevator buttons) than the underlying SARS-CoV-2 detection status (positive or negative) across all variables examined. This is despite a high case load of SARS-CoV-2 patients during the window of sampling, and SARS-CoV-2 detection signal correlating with the underlying patient load within a Unit [7].

Indicator species analysis based on SARS-CoV-2 detection identified a handful of strongly correlated microbes, including several soil-associated lineages and three human-microbiome-associated opportunistic pathogens. Only one virus was identified as an indicator from this analysis, an unclassified virus from an unclassified phylum – estimating the host of this virus was not possible. These data suggest that, while the overall microbial community is defined by the environment and other external forces, some organisms are coincident with SARS-CoV-2. These microbes may serve as potential targets to explore microbial community disruption during SARS-CoV-2 infection, either at the host/human level, or in the built environment (e.g., if these lineages more readily withstand enhanced cleaning regimens associated with outbreak wards and the ICU).

Antimicrobial resistance patterns were similarly governed by sample source rather than SARS-CoV-2 detection status, with higher prevalence of resistance mechanisms targeting tetracycline, aminoglycoside, and fluoroquinolone antibiotics, with a strong signal for resistance to multiple drug classes. Key ARGs of interest were identified based on clinical relevance – these showed four broad categories of abundance, from consistently very low or low detection levels to consistently high. One ARG, *vanA*, showed higher variability in its abundance across Units and a statistically significant difference between its detection on floors versus elevator buttons. *vanA* is part of a gene cluster that is responsible for high-level vancomycin resistance in *Enterococcus* species (such as *Enterococcus faecium* and *Enterococcus faecalis*). Vancomycin resistant *Enterococcus* (VRE) are considered environmentally entrenched in hospitals - the presence of *vanA* was expected, but its variable prevalence across units and time (Figure 5) indicates built environment swabbing may allow identification of target areas to reduce VRE in hospitals [36]. This study identified key microbial community members associated with SARS-CoV-2 detection on hospital built environment surfaces, and demonstrated the ability to detect both punctuated viral signatures and varying ARG prevalence. Future work will determine the correlations between this detection capacity and the associated disease and/or resistance burden within the hospital to explore the potential of built environmental surveillance as an additional mechanism for monitoring outbreaks and other health risks developing in a unit or over time. SARS-CoV-2 co-associated organisms are interesting targets to explore within a human microbiome survey, to identify whether these lineages are associated with COVID-19 infection dynamics, severity, or long-term impacts on host health. An important limitation to our study is that we lacked human-level clinical data. For example, in the location where we found a high abundance of herpesvirales we don’t know whether there was a patient hospitalized with a related infection at that point in time. Future studies that include not only built environment data but also clinical data are needed to better contextualize and understand the associations we observed in our study.

## Declarations

### Ethics approval and consent to participate

Not applicable.

## Consent for publication

Not applicable.

## Availability of data, materials, and software

All human-subtracted read datasets are publicly available from the SRA database under Biosample accessions SAMN46426143 - SAMN46426180.

## Authors’ contributions

NAG: Data Curation; Formal Analysis; Methodology; Visualization; Writing – Original Draft Preparation; Writing – Review & Editing

LB: Data Curation; Formal Analysis; Methodology; Validation; Visualization; Writing – Review & Editing

AH: Data Curation; Formal Analysis; Methodology; Writing – Review & Editing MEK: Data Curation; Formal Analysis Lydia Xing: Investigation

Evgueni Doukhanine: Conceptualization; Funding Acquisition; Methodology; Writing – Review & Editing

DRMacF: Conceptualization; Funding Acquisition; Methodology; Project Administration; Resources; Supervision; Writing – Review & Editing

CN: Conceptualization; Funding Acquisition; Methodology; Project Administration; Writing – Review & Editing

MF: Conceptualization; Funding Acquisition; Methodology; Project Administration; Writing – Review & Editing

RK: Conceptualization; Funding Acquisition; Methodology; Project Administration; Supervision; Writing – Review & Editing

AW: Conceptualization; Funding Acquisition; Methodology; Project Administration; Resources; Software; Supervision; Writing – Review & Editing

LAH: Conceptualization; Data Curation; Formal Analysis; Funding Acquisition; Investigation; Methodology; Project Administration; Resources; Software; Supervision; Validation; Visualization; Writing – Original Draft Preparation; Writing – Review & Editing

## Acknowledgements

We thank Dr. Morgan Langille’s lab for providing us advanced access to the NCBI RefSeq Complete V205 500GB Kraken2 database.

